# Association of Toll-like receptor 7 variants with life-threatening COVID-19 disease in males

**DOI:** 10.1101/2020.11.19.20234237

**Authors:** Chiara Fallerini, Sergio Daga, Stefania Mantovani, Elisa Benetti, Aurora Pujol, Nicola Picchiotti, Agatha Schluter, Laura Planas-Serra, Jesús Troya, Margherita Baldassarri, Francesca Fava, Serena Ludovisi, Francesco Castelli, Maria Eugenia Quiros-Roldan, Massimo Vaghi, Stefano Rusconi, Matteo Siano, Maria Bandini, Simone Furini, Francesca Mari, GEN-COVID Multicenter Study, Alessandra Renieri, Mario U. Mondelli, Elisa Frullanti

## Abstract

**Background:** COVID-19 clinical presentation ranges from asymptomatic to fatal outcome. This variability is due in part to host genome specific mutations. Recently, two families in which COVID-19 segregates like an X-linked recessive monogenic disorder environmentally conditioned by SARS-CoV-2 have been reported leading to identification of loss-of-function variants in *TLR7*.

**Objective:** We sought to determine whether the two families represent the tip of the iceberg of a subset of COVID-19 male patients.

**Methods:** We compared male subjects with extreme phenotype selected from the Italian GEN-COVID cohort of 1178 SARS-CoV-2-infected subjects (<60y, 79 severe cases versus 77 control cases). We applied the LASSO Logistic Regression analysis, considering only rare variants on the young male subset, picking up *TLR7* as the most important susceptibility gene.

**Results:** Rare *TLR7* missense variants were predicted to impact on protein function in severely affected males and in none of the asymptomatic subjects. We then investigated a similar white European cohort in Spain, confirming the impact of *TRL7* variants. A gene expression profile analysis in peripheral blood mononuclear cells after stimulation with TLR7 agonist demonstrated a reduction of mRNA level of TLR7, IRF7, ISG15, IFN-□ and IFN-γ in COVID-19 patients compared with unaffected controls demonstrating an impairment in type I and II INF responses.

**Conclusion:** Young males with *TLR7* loss-of-function mutations and severe COVID-19 in the two reported families represent only a fraction of a broader and complex host genome situation. Specifically, missense mutations in the X-linked recessive *TLR7* disorder may significantly contribute to disease susceptibility in up to 4% of severe COVID-19.

**Clinical Implication:** In this new yet complex scenario, our observations provide the basis for a personalized interferon-based therapy in patients with rare *TLR7* variants.

**CAPSULE SUMMARY:** Our results in large cohorts from Italy and Spain showed that X-linked recessive TLR7 disorder may represent the cause of disease susceptibility to COVID-19 in up to 4% of severely affected young male cases.

## INTRODUCTION

Coronavirus disease 2019 (COVID-19), a potentially severe systemic disease caused by coronavirus SARS-CoV-2, is characterized by a highly heterogeneous phenotypic variability, with the large majority of infected individuals experiencing only mild or even no symptoms. However, severe cases can rapidly evolve towards a critical respiratory distress syndrome and multiple organ failure^1^. COVID-19 still represents an enormous challenge for the world’s healthcare systems ten months after the first appearance in December 2019 in Wuhan, Huanan, Hubei Province of China. Symptoms of COVID-19 range from fever, cough, sore throat, congestion, anosmia and fatigue to shortness of breath, interstitial pneumonia followed by respiratory distress and septic shock^1^. Although older age and the presence of cardiovascular or metabolic comorbidities have been identified as risk factors predisposing to severe disease^2^, these factors alone do not fully explain the differences in the severity. Male patients show more severe clinical manifestations than females with higher prevalence of hospitalizations (16% versus 12%), ICU admissions (3% versus 2%), and deaths (6% versus 5%), indicating that gender may influence the outcome of the disease^3^. These findings suggest a role of host predisposing genetic factors in the pathogenesis of the disease which may be responsible for different clinical outcomes as a result of different antiviral defence mechanisms as well as specific receptor permissiveness to virus and immunogenicity.

More than that, recent evidences suggest also a fundamental role of IFN genes in genetic bases of COVID-19; in particular, recently rare variants have been identified in the interferon type I pathway that are responsible for inborn errors of immunity in a small percentage of patients and auto-antibodies against type I interferon genes in up to 10% of severe COVID-19 cases^4,5^.

Toll-like receptors (TLRs) are crucial components in the initiation of innate immune response - the non specific-immune response-to a variety of pathogens, causing the production of pro-inflammatory cytokines (TNF-α, IL-1, and IL-6) and type I and II IFNs, that are responsible for innate antiviral responses. In particular the innate immune response is very sensitive in detecting potentially pathogenic antigens, activating downstream signaling to induce transcription factors in the nucleus, promoting synthesis and release of types I and type III interferons (IFNs) in addition to a number of other important proinflammatory cytokines. However, when the immune response is dysregulated, it can result in excessive inflammation, or in extreme cases, death. Among them, TLR7 recognises several single-stranded RNA viruses including SARS-CoV-2^6^. We previously showed that another RNA virus, hepatitis C virus (HCV), is able to inhibit CD4 T cell function via Toll-like receptor 7 (TLR7)^7^. Recently, van der Made *et al*. have reported two independent families in which COVID-19 segregates like an X-linked recessive monogenic disorder conditioned by SARS-CoV-2, as an environmental factor^8^.

Here, we performed a nested case-control study (NCC) within our prospectively recruited GEN-COVID cohort with the aim to determine whether the 2 families described by van der Made *et al*. represent an ultra-rare situation or the tip of the iceberg of a larger subset of young male patients.

## RESULTS AND DISCUSSION

We applied LASSO logistic regression analysis, after correcting for Principal Components, to a synthetic boolean representation of the entire set of genes of the X chromosome on the extreme phenotypic ends of the young male subset of the Italian GEN-COVID cohort (https://sites.google.com/dbm.unisi.it/gen-covid)^9^. Cases were selected according to the following inclusion criteria: *i*. male gender; *ii*. young age (<60 years); *iii* endotracheal intubation or CPAP/biPAP ventilation (79 subjects). As controls, 77 subjects were selected using the sole criterion of being oligo-asymptomatic not requiring hospitalization. Cases and controls represented the extreme phenotypic presentations of the GEN-COVID cohort. Exclusion criteria for both cases and controls were: *i*. SARS-CoV-2 infection not confirmed by PCR; *ii*. non-caucasian ethnicity. The materials and methods details are listed in the Online Repository section.

The study (GEN-COVID) was consistent with Institutional guidelines and approved by the University Hospital (Azienda Ospedaliero-Universitaria Senese) Ethical Review Board, Siena, Italy (Prot n. 16929, dated March 16, 2020). Only rare variants (≤1% in European Non-Finnish population) were considered in the boolean representation: the gene was set to 1 if it included at least a missense, splicing, or loss of function rare variant, and 0 otherwise. Fisher Exact test was then used for the specific data validation.

Toll-like receptor 7 (*TLR7*) was picked up as one of the most important susceptibility genes by LASSO Logistic Regression analysis (**Figure 1**). We then queried the COVID-19 section of the Network of Italian Genome NIG database (http://www.nig.cineca.it/, specifically, http://nigdb.cineca.it) that houses the entire GEN-COVID cohort represented by more than 1000 WES data of COVID-19 patients and SARS-CoV-2 infected asymptomatic subjects^7^. By selecting for young (<60 year-old) males, we obtained rare (MAF≤1%) *TLR7* missense variants predicted to impact on protein function (CADD>12.28) in 5 out of 79 male patients (6,32%) with life-threatening COVID-19 (hospitalized intubated and hospitalized CPAP/BiPAP) and in none of the 77 SARS-CoV2 infected oligo-asymptomatic male subjects.

**Figure 1.**
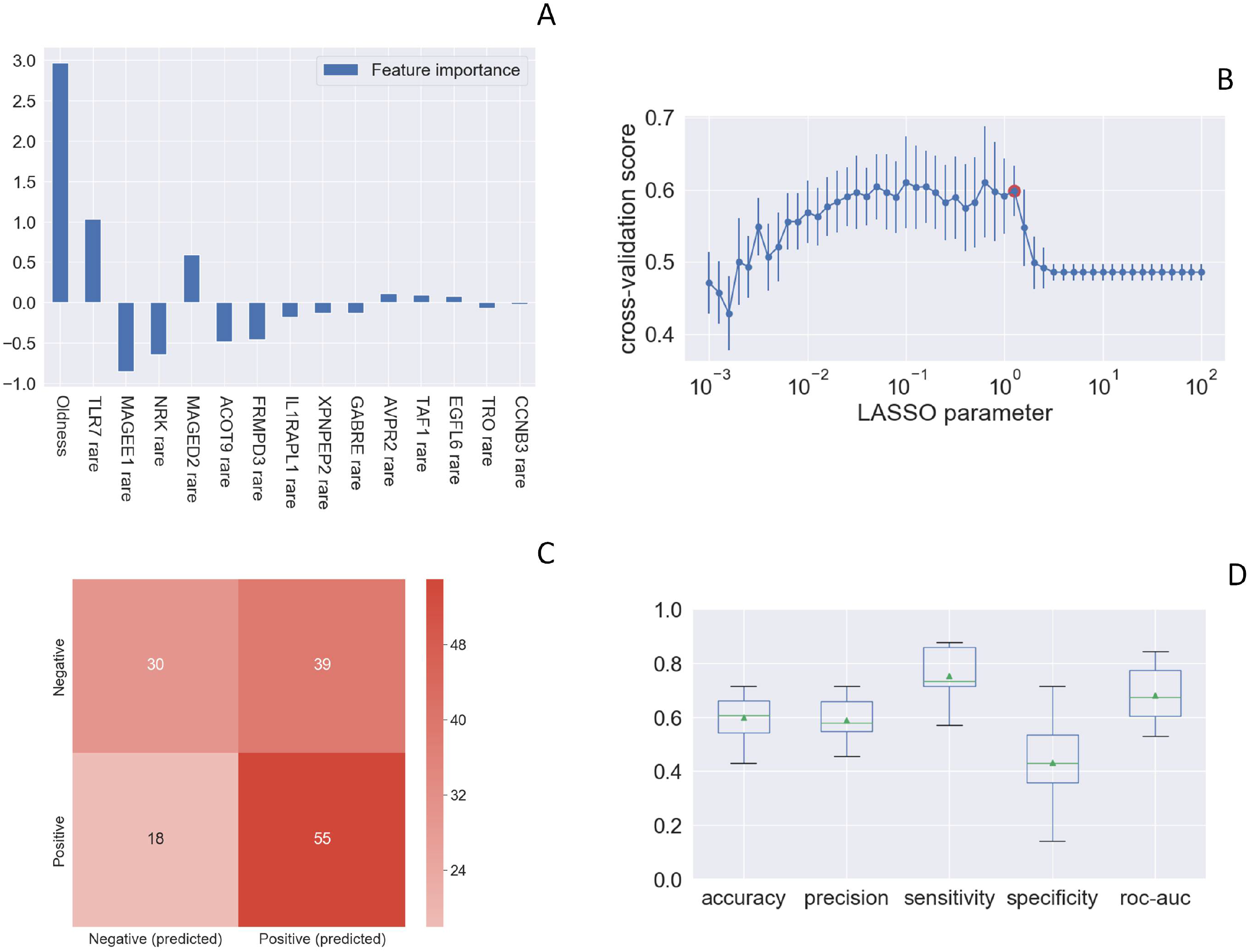
Rare *TLR7* variants and association with COVID-19. LASSO logistic regression on boolean representation of rare variants of all genes of the X chromosome (X chr). TLR7 is picked up by LASSO logistic regression as one of the most important genes on the X chr (**Panel A**). The LASSO logistic regression model provides an embedded feature selection method within the binary classification tasks (male patients with life-threatening COVID-19 vs infected asymptomatic male subjects). The upward histograms (positive weights) reflect a susceptible behaviour of the features to the target COVID-19, whereas the downward histograms (negative weights) a protective action. In addition to the *TLR7* gene, other genes are picked up, including XPNPEP2 dysregulation bradykinine system. **Panel B** represents the cross-validation accuracy score for the grid of LASSO regularization parameters; the error bar is given by the standard deviation of the score within the 10 folds; the red point (1.26) corresponds to the parameter chosen for the fitting procedure. Performances are evaluated through the confusion matrix of the aggregated predictions in the 10 folds of the cross-validation (**Panel C**) and with the boxplot (**Panel D**) of accuracy (60% average value), precision (59%), sensitivity (75%), specificity (43%), and ROC-AUC score (68%). The box extends from the Q1 to Q3 quartile, with a line at the median (Q2) and a triangle for the average.

A similar Spanish cohort, composed of male patients under 60 years of age without comorbidities (77 cases and 45 controls) was used to expand the cohort to another representative European population highly impacted by COVID-19. All subjects were white European. The Spanish Covid HGE cohort is under IRB approval PR127/20 from Bellvitge University Hospital, Barcelona, Spain. We found a *TLR7* variant in one of 77 cases (1,29%) and in none of 45 controls. Overall, the association between the presence of *TLR7* rare mutations and severe COVID-19 was significant (p= 0.04 by Fisher Exact test, **Table 1**).

**Table 1.** Fisher exact test of the overall combined cohorts in young males (<60 years).

| Clinical Category | N. Wild Type patients | N. Mutated patients | Total |
| --- | --- | --- | --- |
| Severely Affected Males | 150 | 6 | 156 |
| Asymptomatic Males | 122 | 0 | 122 |
| **Total** | 272 | 6 | 278 (Grand Total) |
| p-value = 0.04 |  |  |  |

We then investigated the presence of *TLR7* mutations in the entire male cohort of COVID-19 (cases and controls) regardless of age. We found *TLR7* rare missense variants in 3 additional patients over 60 years of age, including 2 among cases and 1 among controls, the last bearing the mutation p.Val222Asp predicted to have a low impact on protein function (CADD of 5.36) (**Table 2**).

**Table 2A.** *TLR7* mutations in severely affected Italian and Spanish males -all ages-(cases).

| Nucleotide change | Amino acid change | dbSNP | CADD | ExAC_NFE | Clinical Category* | Age | Cohort | Patient ID |
| --- | --- | --- | --- | --- | --- | --- | --- | --- |
| c.655G>A | Val219Ile | rs149314023 | 12.28 | 0.0003 | 4 | 32 | Italian | P1 |
| c.863C>T | Ala288Val | rs200146658 | 15.37 | 0 | 3 | 57 | Italian | P2 |
| c.901T>C | Ser301Pro | - | 26.4 | N/A | 3 | 46 | Italian | P3 |
| c.1343C>T | Ala448Val | rs5743781 | 13.08 | 0.0047 | 3 | 53 | Italian | P4 |
| c.1343C>T | Ala448Val | rs5743781 | 13.08 | 0.0047 | 3 | 58 | Italian | P5 |
| c.1888C>T | His630Tyr | - | 19.16 | 0 | 4 | 50 | Spanish | P6 |
| c.3094G>A | Ala1032Thr | rs147244662 | 22.3 | 0.0006 | 3 | 65 | Italian | P7 |
| c.3094G>A | Ala1032Thr | rs147244662 | 22.3 | 0.0006 | 3 | 66 | Italian | P8 |

**Table 2B.** *TLR7* mutations in asymptomatic males (controls).

| Nucleotide change | Amino acid change | dbSNP | CADD | ExAC_NFE | Clinical Category | Age | Cohort | Patient ID |
| --- | --- | --- | --- | --- | --- | --- | --- | --- |
| c.665T>A | Val222Asp | rs5590784<br>3 | 5.36 | 0.0047 | 0 | 64 | Italian | C1 |

\*Clinical categories: **4.** Hospitalized and intubated patients; **3.** Hospitalized and CPAP-BiPAP and high-flows oxygen treated; **2.** Hospitalized and treated with conventional oxygen support only; **1.** Hospitalized without respiratory support; **0.** Not hospitalized oligo/asymptomatic individuals

In order to functionally link the presence of the identified *TLR7* missense mutations and the effect on the downstream type I IFN-signaling, we performed a gene expression profile analysis in peripheral blood mononuclear cells (PBMCs) isolated from patients after stimulation with the TLR7 agonist imiquimod (as reported by van der Made *et al*.^8^). PBMCs from 5 out of 8 cases and from the control were available. All *TLR7* variants were tested except one (Ala288Val) as some different patients share the same. This analysis showed a statistically significant decrease of *IRF7* (member of the interferon regulatory transcription factor) and *IFN-*γ mRNA levels in all tested mutations (Val219Ile, Ser301Pro, Ala1032Thr, His630Tyr) identified in cases (P1, P3, P7, P8 and P6) compared with healthy (Ctl) demonstrating a complete impairment in IFN-□and IFN-γ production in response to TLR7 stimulation in all patients (**Figure 2**). A similarly significant reduction was shown for ISG15 (Interferon-stimulated gene 15) and *IFN-*□ mRNA level for 4 out of 5 patients (P3, P8, P6, P7). An analogous trend was observed for TLR7 in P8 and P6 cases. A significant reduction was also shown for all TLR7-related mRNA transcripts when cases were compared with the asymptomatic controls (C1) (**Supplementary Figure 1**). Interestingly, the p.Val222Asp variant proved to be functionally neutral in keeping with its discovery in control and not in cases (**Figure 2**). TLR7 expression was evaluated in monocytes and B cells from patients and healthy controls by flow cytometry. As shown in **Supplementary Figure 2** patients and controls expressed the TLR7 protein at intracellular level. The functional capacity of PBMC was evaluated after stimulation with the TLR4 agonist lipopolysaccharide (LPS). The analysis showed that LPS-induced production of IL6 by CD3-CD14+ cells was similar in patients and controls (**Supplementary Figure 3**).

**Figure 2.**
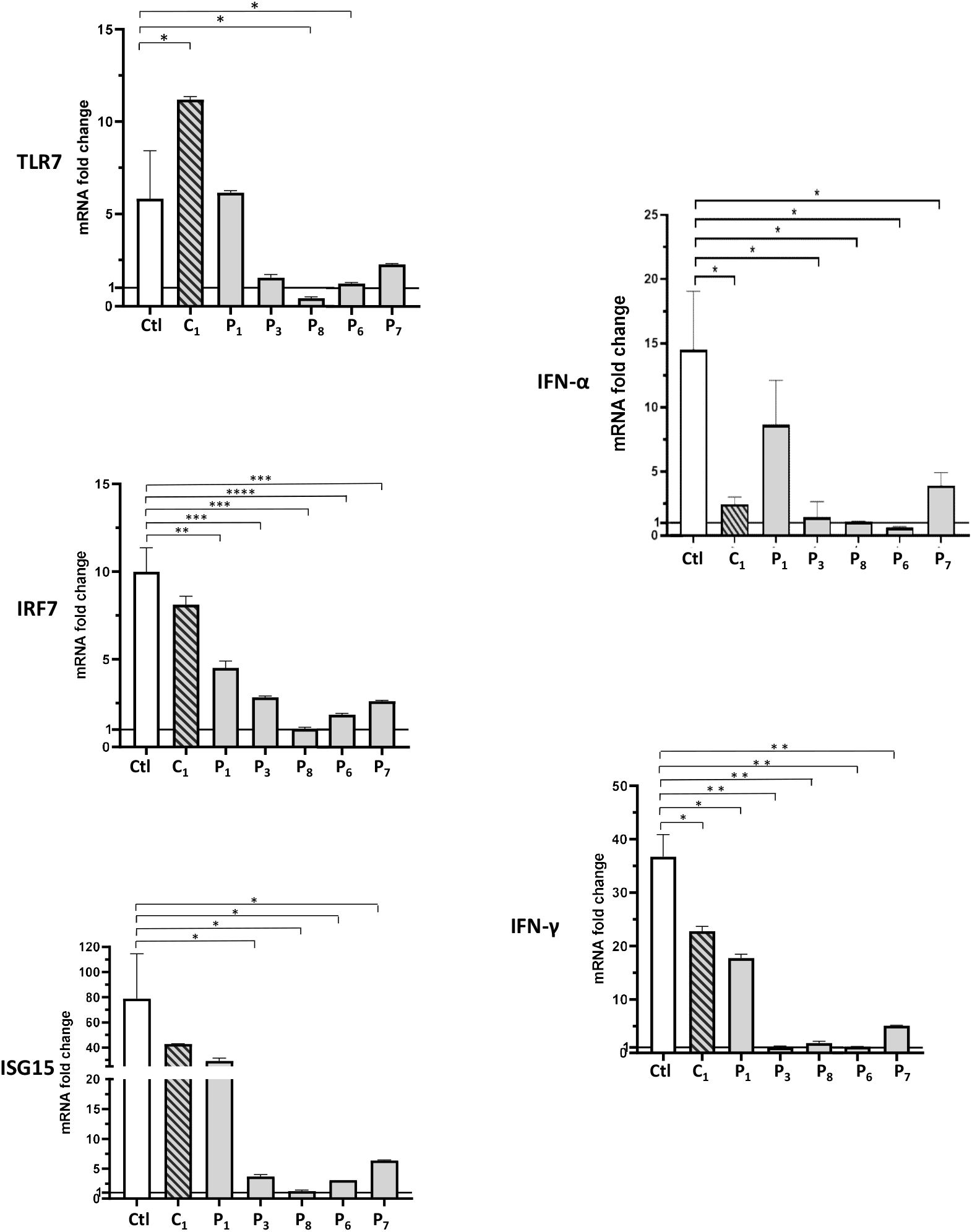
Gene expression profile analysis in peripheral blood mononuclear cells (PBMCs) after stimulation with a TLR7 agonist for 4 hours. 5×10^5^ PBMCs from COVID-19 patients and four unaffected male and female controls were stimulated for 4 hours with the TLR7 agonist imiquimod at 5 μg/mL or cell culture medium. Quantitative PCR assay was performed and the 2^- ΔΔ Ct^ calculated using *HPRT1* as housekeeping gene. Fold change in mRNA expression of *TLR7* and type 1 IFN-related genes *ISG15, IRF7, IFN-*□ and *IFN-*γ induced by TLR7 agonist imiquimod was compared with cell culture medium. Error bars show standard deviations. *P values* were calculated using an unpaired *t* test: *P < .05; **P < .01; ***P < .001; ****P < .0001. Ctl indicates healthy controls; C_1_, the asymptomatic mutated control; P1, P3, P8, P6, P7 severely affected mutated cases (as in **Table 2A**).

**Figure 3.**
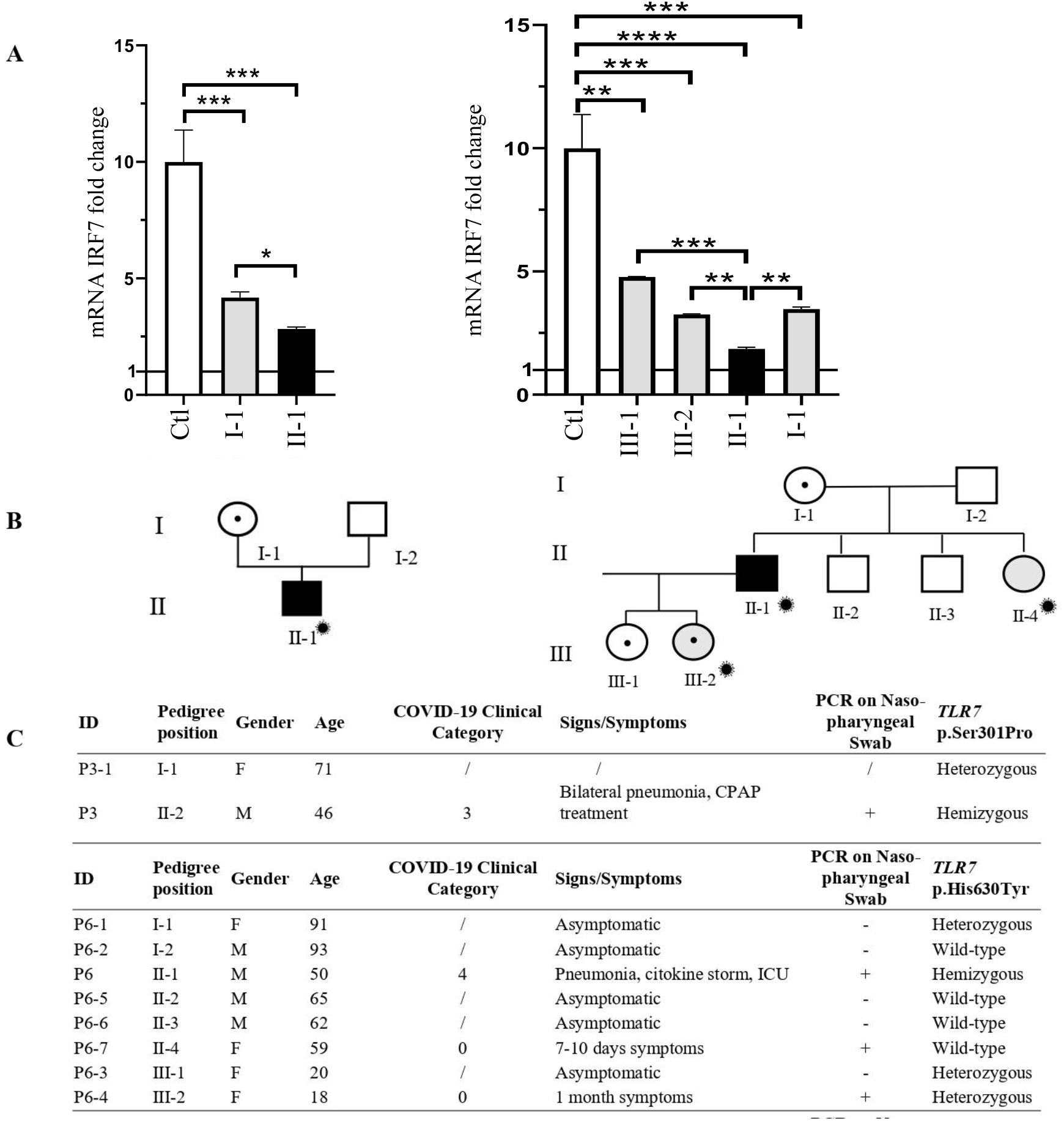
Segregation analysis in Italian (P3) and Spanish family (P6). Fold change in mRNA expression of the main *TLR7*-effector, *IRF7*, is shown in **Panel A**. Pedigree (**Panel B**) and respective segregation of *TLR7* variant and COVID-19 status (**Panel C**). Squares represent male family members; circles, females.

Segregation analysis was available for two of 8 cases, P3 and P6 (**Figure 3**). In both pedigree, the disease nicely segregates as an X-linked disorder conditioned by environmental factors, i.e. SARS-Cov-2 (**Figure 3, panel B**). This was also supported by functional analysis on all *TLR7*-related genes (**Supplementary Figure 4**). As an example, we showed gene expression profile analysis for *IRF7* gene: in male patients *IRF7* mRNA levels are statistically significantly reduced compared to heterozygous females (**Figure 3, panel A**). Of note, only the infected mutated male subject had severe COVID-19, whereas the infected not mutated female (sister) was oligosymptomatic with symptoms lasting 7-10 days; the infected mutated female (one of the daughters) was oligosymptomatic with symptoms lasting one month (**Figure 3, panel C**). Both females did not require any hospital treatment.

Our results showed that the two families with loss of function mutations reported by van der Made *et a*l.^8^ in males with severe COVID-19 with a mean age of 26 years are indeed the tip of the iceberg of a broader host genome scenario. Specifically, missense mutations in the X-linked recessive TLR7 disorder may represent the cause of disease susceptibility to COVID-19 in up to 4% of severely affected young male cases (6/156, 3,84%). Since not all identified mutations were functionally tested, the true percentage could be slightly lower. Overall, males with missense mutations shown here developed COVID-19 at a mean age of 58 years, considerably later than 26 years, in agreement with a predicted smaller impact on the protein than the loss of function mutations reported by van der Made *et al*.^8^. Similarly, the identified rare missense *TLR7* variants impaired the mRNA expression of *TLR7* as well as the downstream pathway. The observation reported here may lead to consider *TLR7* screening in severely affected male patients in order to start personalized interferon treatment for those with this specific genetic disorder.

## Supporting information

Online Repository

## Data Availability

The data and samples referenced here are housed in the GEN-COVID Patient Registry and the GEN-COVID Biobank and are available for consultation. For further information, you may contact the corresponding author, Prof. Alessandra Renieri.

http://www.nig.cineca.it

https://www.covid19hg.org/

http://nigdb.cineca.it

## ACKNOWLEDGEMENTS

This study is part of the GEN-COVID Multicenter Study, https://sites.google.com/dbm.unisi.it/gen-covid, the Italian multicenter study aimed at identifying the COVID-19 host genetic bases. Specimens were provided by the COVID-19 Biobank of Siena, which is part of the Genetic Biobank of Siena, member of BBMRI-IT, of Telethon Network of Genetic Biobanks (project no. GTB18001), of EuroBioBank, and of D-Connect. We thank the CINECA consortium for providing computational resources and the Network for Italian Genomes (NIG) http://www.nig.cineca.it for its support. We thank private donors for the support provided to A.R. (Department of Medical Biotechnologies, University of Siena) for the COVID-19 host genetics research project (D.L n.18 of March 17, 2020). We also thank the COVID-19 Host Genetics Initiative (https://www.covid19hg.org/). MIUR project “Dipartimenti di Eccellenza 2018-2020” to the Department of Medical Biotechnologies University of Siena, Italy. Research on the Spanish cohort received funding from the European Union’s Horizon 2020 research and innovation programme under grant agreement No 824110 – EASI-Genomics, to AP.

## Abbreviations

Ala: Alanine amino acid
Asp: Asparagine amino acid
CADD: Combined Annotation Dependent Depletion
COVID-19: COronaVIrus Disease 19
His: Histidine amino acid
HPRT1: hypoxanthine phosphoribosyltransferase 1
INFs: Interferons
IFN-α: Interferon Alpha
IFN-β1: Interferon Beta1
IFN-γ: Interferon Gamma
IL-1: Interleukin 1
IL-6: Interleukin 6
Ile: Isoleucine amino acid
IRF7: Interferon Regulatory Factor 7
ISG15: Interferon-Stimulated Gene 15
LASSO: Least Absolute Shrinkage and Selection Operator
LBP: LipoPolySaccharide
(LPS): Binding Protein
NIG: Network of Italian Genome
PBMCs: Peripheral Blood Mononuclear Cells
Pro: Proline amino acid
RNA: RiboNucleic Acid
Ser: Serine amino acid
SARS-CoV-2: Severe Acute Respiratory Syndrome caused by CoronaVirus
Thr: Threonine amino acid
TLRs: Toll-like Receptors
TLR4: Toll-like Receptor 4
TLR7: Toll-like Receptor 7
TNF-α: Tumor Necrosis Factor α
Tyr: Tyrosine amino acid
Val: Valine amino acid
WES: Whole Exome Sequencing

## ADDITIONAL INFORMATION

### GEN-COVID Multicenter Study (https://sites.google.com/dbm.unisi.it/gen-covid)

Floriana Valentino^1,2^, Gabriella Doddato^1,2^, Annarita Giliberti^1,2^, Rossella Tita^9^, Sara Amitrano^9^, Mirella Bruttini^1,2,9^, Susanna Croci^1,2^, Ilaria Meloni^1,2^, Maria Antonietta Mencarelli^9^, Caterina Lo Rizzo^9^, Anna Maria Pinto^9^, Laura Di Sarno^1,2^, Giada Beligni^1,2^, Andrea Tommasi^1,2,9^, Maria Palmieri^1,2^, Francesca Montagnani^2,17^, Arianna Emiliozzi^2,17^, Massimiliano Fabbiani^17^, Barbara Rossetti^17^, Giacomo Zanelli^2,17^, Elena Bargagli^18^, Laura Bergantini^18^, Miriana D’Alessandro^18^, Paolo Cameli^18^, David Bennet^18^, Federico Anedda^19^, Simona Marcantonio^19^, Sabino Scolletta^19^, Federico Franchi^19^, Maria Antonietta Mazzei^20^, Susanna Guerrini^20^, Edoardo Conticini^21^, Luca Cantarini^21^, Bruno Frediani^21^, Danilo Tacconi^22^, Chiara Spertilli^22^, Marco Feri^23^, Alice Donati^23^, Raffaele Scala^24^, Luca Guidelli^24^, Genni Spargi^25^, Marta Corridi^25^, Cesira Nencioni^26^, Leonardo Croci^26^, Gian Piero Caldarelli^27^, Maurizio Spagnesi^16^, Paolo Piacentini^16^, Elena Desanctis^16^, Silvia Cappelli^16^, Anna Canaccini^28^, Agnese Verzuri^28^, Valentina Anemoli^28^, Agostino Ognibene^29^, Antonella D’Arminio Monforte^30^, Esther Merlini^30^, Federica Gaia Miraglia^30^, Massimo Girardis^31^, Sophie Venturelli^31^, Marco Sita^31^, Andrea Cossarizza^32^, Andrea Antinori^33^, Alessandra Vergori^33^, Arianna Gabrieli^15^, Agostino Riva^14,15^, Daniela Francisci^34,35^, Elisabetta Schiaroli^34^, Pier Giorgio Scotton^36^, Francesca Andretta^36^, Sandro Panese^37^, Renzo Scaggiante^38^, Francesca Gatti^38^, Saverio Giuseppe Parisi^39^, Stefano Baratti^39^, Melania Degli Antoni^12^, Isabella Zanella^12^, Matteo Della Monica^40^, Carmelo Piscopo^40^, Mario Capasso^41,42,43^, Roberta Russo^41,42^, Immacolata Andolfo^41,42^, Achille Iolascon^41,42^, Giuseppe Fiorentino^44^, Massimo Carella^45^, Marco Castori^45^, Giuseppe Merla^45^, Filippo Aucella^46^, Pamela Raggi^47^, Carmen Marciano^47^, Rita Perna^47^, Matteo Bassetti^48,49^, Antonio Di Biagio^49^, Maurizio Sanguinetti^50,51^, Luca Masucci^50,51^, Serafina Valente^52^, Marco Mandalà^53^, Alessia Giorli^53^, Lorenzo Salerni^53^, Patrizia Zucchi^54^, Pierpaolo Parravicini^54^, Elisabetta Menatti^55^, Tullio Trotta^56^, Ferdinando Giannattasio^56^, Gabriella Coiro^56^, Fabio Lena^57^, Domenico A. Coviello^58^, Cristina Mussini^59^, Giancarlo Bosio^60^, Enrico Martinelli^60^, Sandro Mancarella^61^, Luisa Tavecchia^61^, Marco Gori^8,6^, Lia Crotti^63,64,65,66^, Agatha Schluter^4,5,67^, Laura Planas-Serra^4,5,67^, Marta Gut^68^, Chiara Gabbi^69^.

17) Dept of Specialized and Internal Medicine, Tropical and Infectious Diseases Unit, Azienda Ospedaliera Universitaria Senese, Siena, Italy

18) Unit of Respiratory Diseases and Lung Transplantation, Department of Internal and Specialist Medicine, University of Siena, Italy

19) Dept of Emergency and Urgency, Medicine, Surgery and Neurosciences, Unit of Intensive Care Medicine, Siena University Hospital, Italy

20) Department of Medical, Surgical and Neuro Sciences and Radiological Sciences, Unit of Diagnostic Imaging, University of Siena, Italy

21) Rheumatology Unit, Department of Medicine, Surgery and Neurosciences, University of Siena, Policlinico Le Scotte, Italy

22) Department of Specialized and Internal Medicine, Infectious Diseases Unit, San Donato Hospital Arezzo, Italy

23) Dept of Emergency, Anesthesia Unit, San Donato Hospital, Arezzo, Italy

24) Department of Specialized and Internal Medicine, Pneumology Unit and UTIP, San Donato Hospital, Arezzo, Italy

25) Department of Emergency, Anesthesia Unit, Misericordia Hospital, Grosseto, Italy

26) Department of Specialized and Internal Medicine, Infectious Diseases Unit, Misericordia Hospital, Grosseto, Italy

27) Clinical Chemical Analysis Laboratory, Misericordia Hospital, Grosseto, Italy

28) Territorial Scientific Technician Department, Azienda USL Toscana Sud Est, Italy

29) Clinical Chemical Analysis Laboratory, San Donato Hospital, Arezzo, Italy

30) Department of Health Sciences, Clinic of Infectious Diseases, ASST Santi Paolo e Carlo, University of Milan, Italy

31) Department of Anesthesia and Intensive Care, University of Modena and Reggio Emilia, Modena, Italy

32) Department of Medical and Surgical Sciences for Children and Adults, University of Modena and Reggio Emilia, Modena, Italy

33) HIV/AIDS Department, National Institute for Infectious Diseases, IRCCS, Lazzaro Spallanzani, Rome, Italy

34) Infectious Diseases Clinic, Department of Medicine 2, Azienda Ospedaliera di Perugia and University of Perugia, Santa Maria Hospital, Perugia, Italy

35) Infectious Diseases Clinic, “Santa Maria” Hospital, University of Perugia, Perugia, Italy

36) Department of Infectious Diseases, Treviso Hospital, Local Health Unit 2 Marca Trevigiana, Treviso, Italy

37) Clinical Infectious Diseases, Mestre Hospital, Venezia, Italy

38) Infectious Diseases Clinic, ULSS1, Belluno, Italy

39) Department of Molecular Medicine, University of Padova, Italy

40) Medical Genetics and Laboratory of Medical Genetics Unit, A.O.R.N. “Antonio Cardarelli”, Naples, Italy

41) Department of Molecular Medicine and Medical Biotechnology, University of Naples Federico II, Naples, Italy

42) CEINGE Biotecnologie Avanzate, Naples, Italy

43) IRCCS SDN, Naples, Italy

44) Unit of Respiratory Physiopathology, AORN dei Colli, Monaldi Hospital, Naples, Italy

45) Division of Medical Genetics, Fondazione IRCCS Casa Sollievo della Sofferenza Hospital, San Giovanni Rotondo, Italy

46) Department of Medical Sciences, Fondazione IRCCS Casa Sollievo della Sofferenza Hospital, San Giovanni Rotondo, Italy

47) Clinical Trial Office, Fondazione IRCCS Casa Sollievo della Sofferenza Hospital, San Giovanni Rotondo, Italy

48) Department of Health Sciences, University of Genova, Genova, Italy

49) Infectious Diseases Clinic, Policlinico San Martino Hospital, IRCCS for Cancer Research Genova, Italy

50) Microbiology, Fondazione Policlinico Universitario Agostino Gemelli IRCCS, Catholic University of Medicine, Rome, Italy

51) Department of Laboratory Sciences and Infectious Diseases, Fondazione Policlinico Universitario A. Gemelli IRCCS, Rome, Italy

52) Department of Cardiovascular Diseases, University of Siena, Siena, Italy

53) Otolaryngology Unit, University of Siena, Italy

54) Department of Internal Medicine, ASST Valtellina e Alto Lario, Sondrio, Italy

55) Study Coordinator Oncologia Medica e Ufficio Flussi Sondrio, Italy

56) First Aid Department, Luigi Curto Hospital, Polla, Salerno, Italy

57) Local Health Unit-Pharmaceutical Department of Grosseto, Toscana Sud Est Local Health Unit, Grosseto, Italy

58) U.O.C. Laboratorio di Genetica Umana, IRCCS Istituto G. Gaslini, Genova, Italy

59) Infectious Diseases Clinics, University of Modena and Reggio Emilia, Modena, Italy

60) Department of Respiratory Diseases, Azienda Ospedaliera di Cremona, Cremona, Italy

61) U.O.C. Medicina, ASST Nord Milano, Ospedale Bassini, Cinisello Balsamo (MI), Italy

62) Université Côte d’Azur, Inria, CNRS, I3S, Maasai

63) Istituto Auxologico Italiano, IRCCS, Department of Cardiovascular, Neural and Metabolic Sciences, San Luca Hospital, Milan, Italy

64) Department of Medicine and Surgery, University of Milano-Bicocca, Milan, Italy

65) Istituto Auxologico Italiano, IRCCS, Center for Cardiac Arrhythmias of Genetic Origin, Milan, Italy

66) Istituto Auxologico Italiano, IRCCS, Laboratory of Cardiovascular Genetics, Milan, Italy

67) Spanish Covid HGE

68) CNAG-CRG, Centre for Genomic Regulation (CRG), Barcelona Institute of Science and Technology (BIST), Carrer Baldiri i Reixac 4, 08028, Barcelona, Spain.

69) Independent Scientist, Milan, Italy

