## Supplementary Figure 1 for "Association of Toll-like receptor 7 variants with life-threatening COVID-19 disease in males"

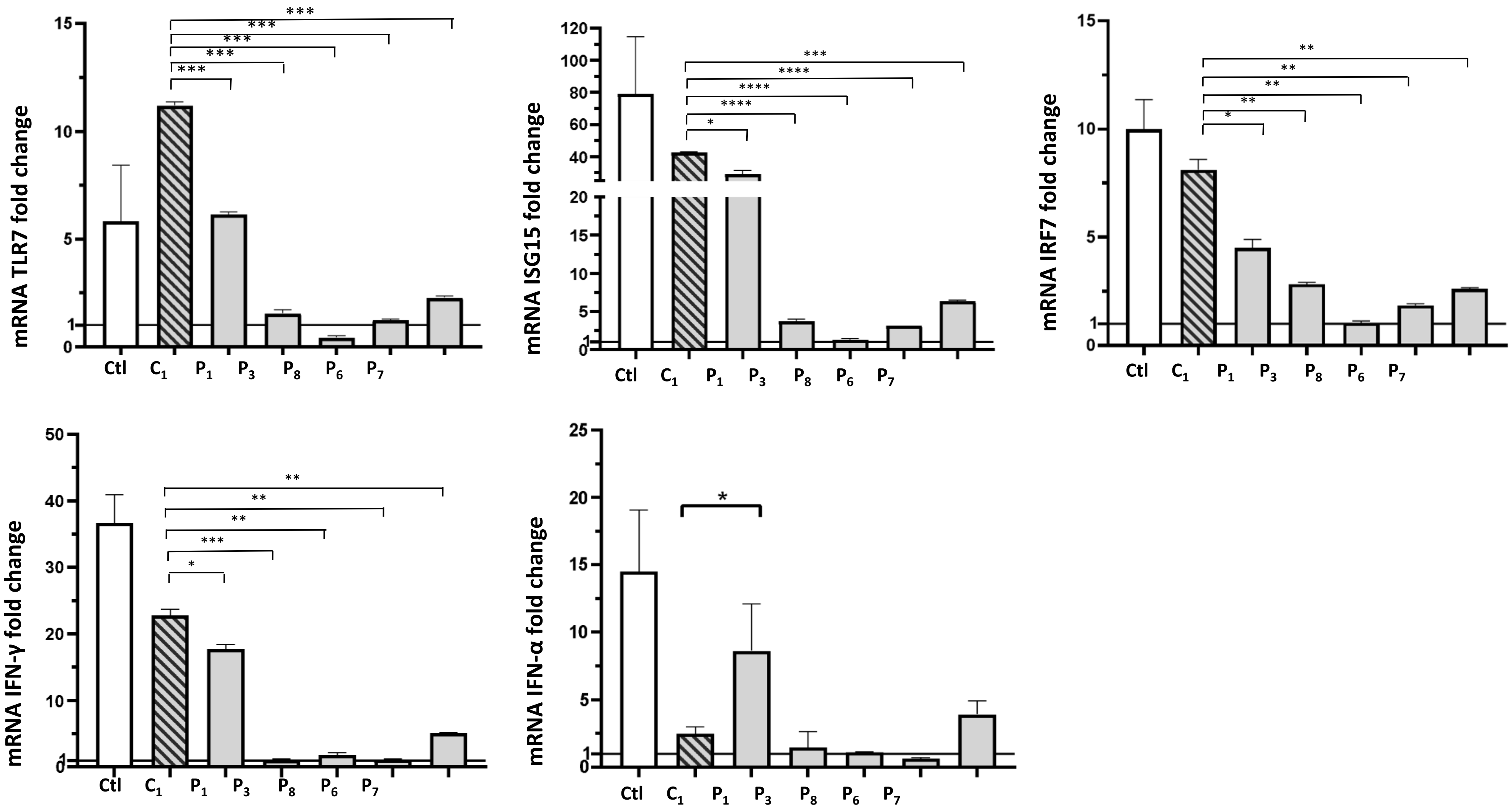


**Supplementary Figure 1.** Gene expression profile analysis in peripheral blood mononuclear cells (PBMCs) after stimulation with a TLR7 agonist for 4 hours in COVID-19 cases compared with the asymptomatic controls (C1). *P values* were calculated using an unpaired *t* test: *P < .05; **P < .01; ***P < .001; ****P < .0001.
