## Supplementary Figure 2 for "Association of Toll-like receptor 7 variants with life-threatening COVID-19 disease in males"

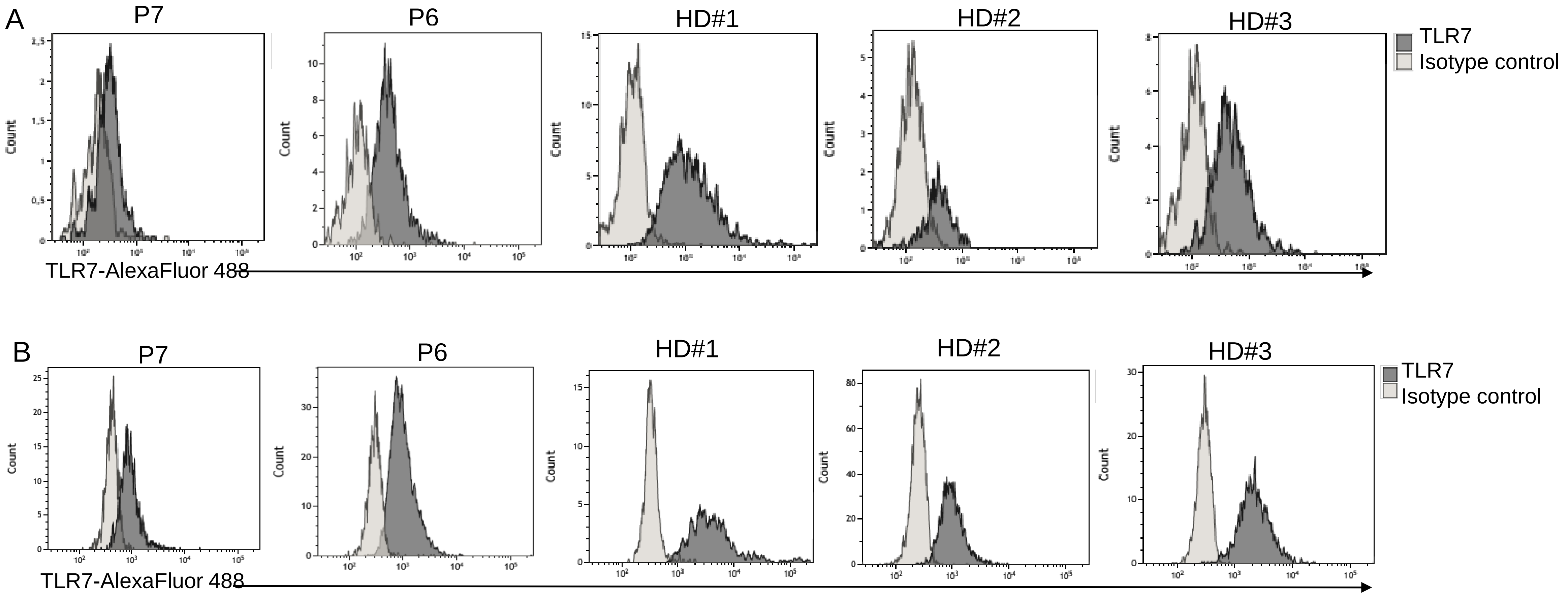


**Supplementary Figure 2.** Representative flow cytometry analysis of intracellularly expressed TLR7 in CD3-CD19+ B cells (A) and in CD3-CD14+ monocyte cells (B) in controls and patients.
