## Supplementary Figure 3 for "Association of Toll-like receptor 7 variants with life-threatening COVID-19 disease in males"

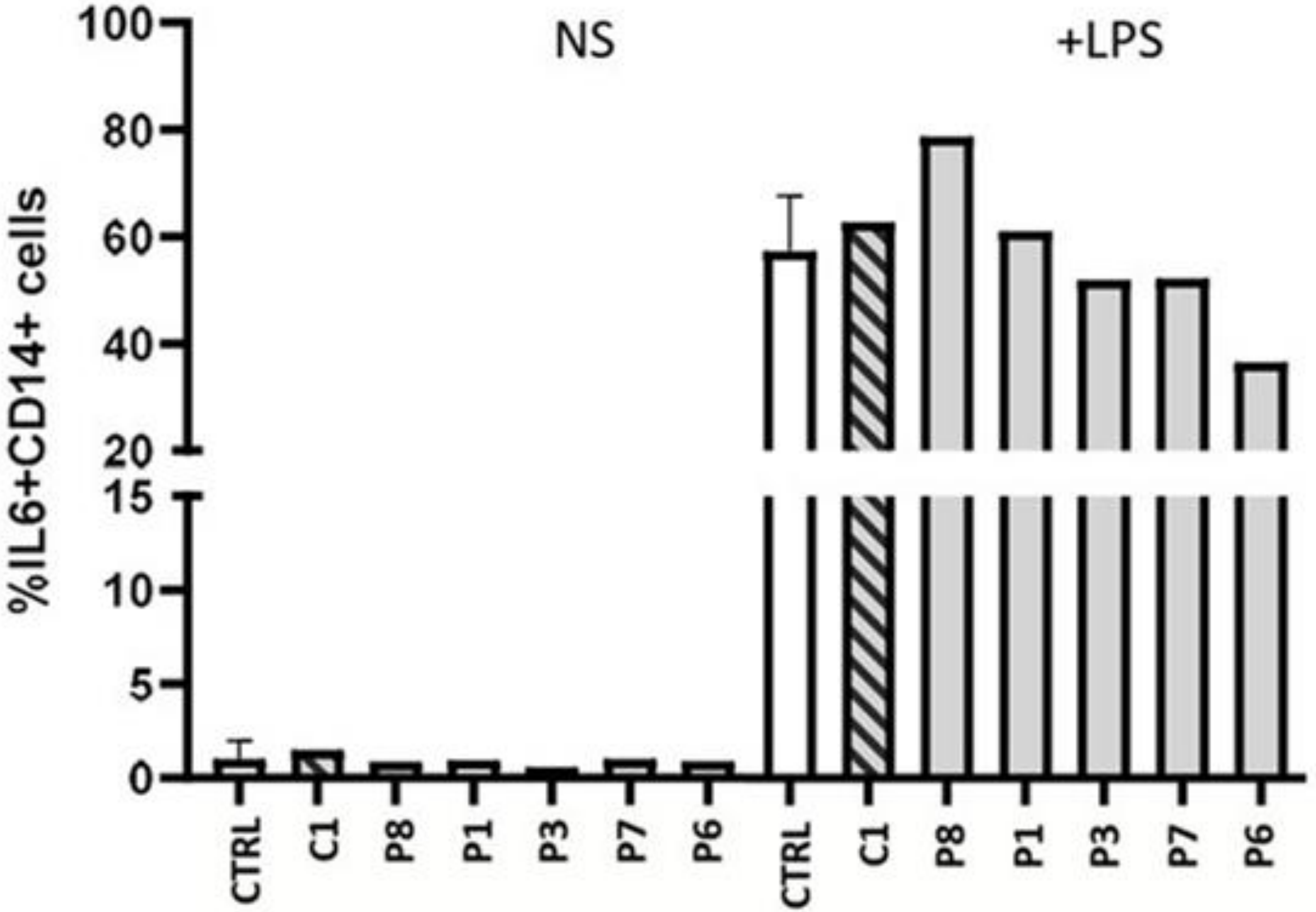


**Supplementary Figure 3.** LPS-induced production of IL6 by CD3-CD14$^{+}$ cells. PBMCs from patients and healthy controls (n=4) were stimulated in vitro with Lipopolysaccharide (LPS) at 100ng/ml for 6h or with medium alone. The percentage of IL6$^{+}$CD14$^{+}$ cells was evaluated by flow cytometry.
