## Supplementary Figure 4 for "Association of Toll-like receptor 7 variants with life-threatening COVID-19 disease in males"

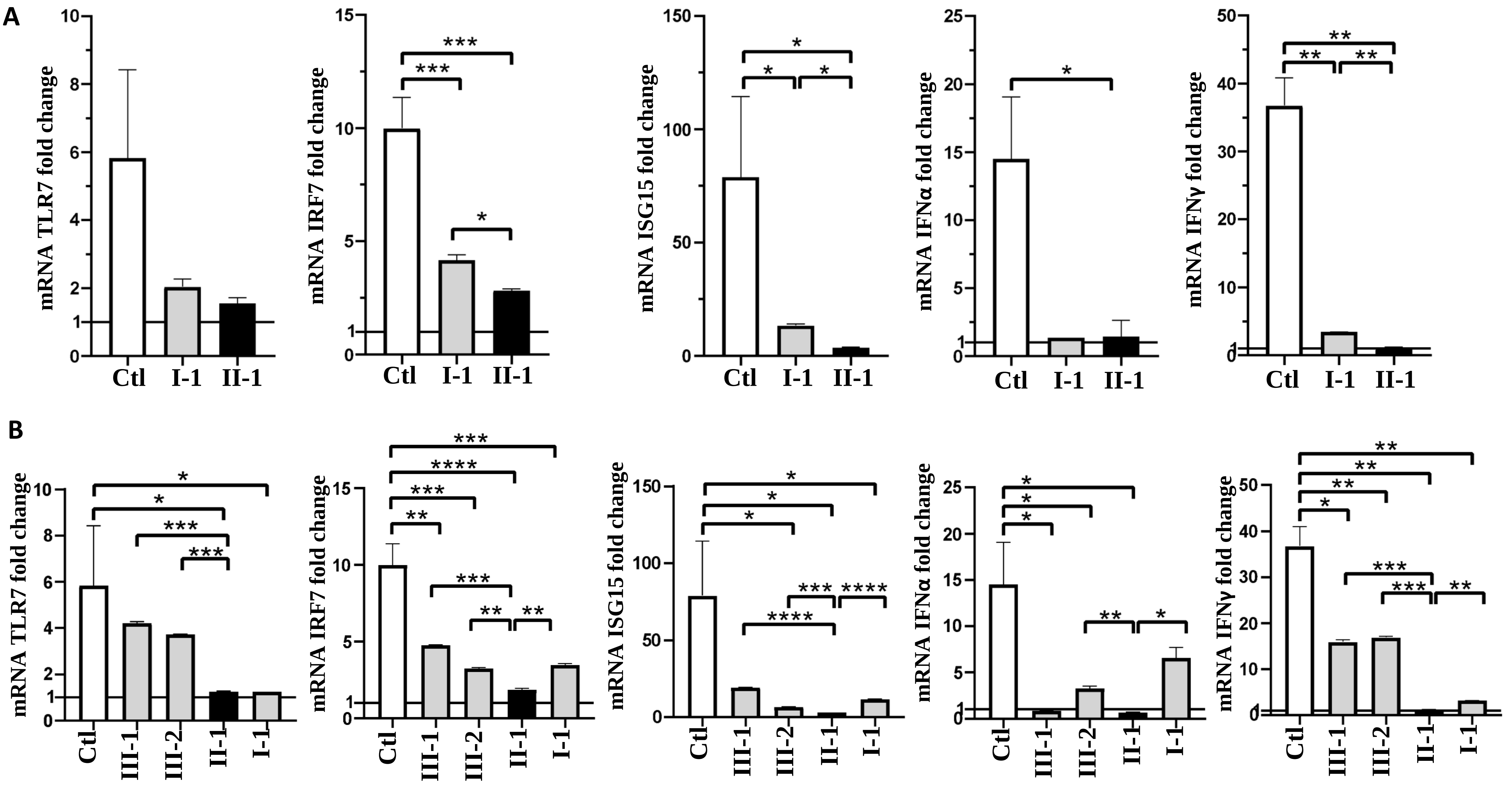


**Supplementary Figure 4.** Gene expression profile analysis in Italian (P3, **Panel A**) and Spanish family (P6, **Panel B**). Fold change in mRNA expression of *TLR7* and type 1 IFN-related genes *ISG15*, *IRF7*, *IFN-a* and *IFN-γ* induced by TLR7 agonist imiquimod. *P values* were calculated using an unpaired *t* test: *P < .05; **P < .01; ***P < .001; ****P < .0001.
