## Supplementary material for "Association of Toll-like receptor 7 variants with life-threatening COVID-19 disease in males": Online Repository

**Patients and samples**

A subset of 156 young (<60 years) male COVID-19 patients was selected from the Italian GEN-COVID cohort of 1178 SARS-CoV-2-infected subjects (<https://sites.google.com/dbm.unisi.it/gen-covid>) ^1^. The study (GEN-COVID) was consistent with Institutional guidelines and approved by the University Hospital (Azienda Ospedaliero-Universitaria Senese) Ethical Review Board, Siena, Italy (Prot n. 16929, dated March 16, 2020). A Spanish cohort, composed of 122 COVID-19 patients, was used to expand the cohort to another representative European population highly impacted by COVID-19. The Spanish Covid HGE cohort is under IRB approval PR127/20 from Bellvitge University Hospital, Barcelona, Spain.

**Statistical Methods**

We adopted the LASSO logistic regression, one of the most common Machine Learning algorithms for classification, that provides a feature selection method within the classification task able to enforce both the sparsity and the interpretability of the results^2^. In fact, the coefficients of the logistic regression model are directly related to the importance of the corresponding features, and LASSO regularization shrinks close to zero the coefficients of features that are not relevant in predicting the response, reducing overfitting.

The Principal components analysis (PCA) was applied prior to the LASSO logistic regression in order to remove samples that were clear outliers with respect to the first 3 principal components from the following analyses (deviating more than 5 standard deviations from the average).

A 10-fold cross-validation method was applied in order to test the performances. It provides the partition of the dataset into 10 batches, then 9 batches are exploited for the training of the LASSO logistic regression and the remaining batch as a test, by repeating this procedure 10 times. The performance metrics are averaged on the 10 testing sets in order to avoid overfitting. The confusion matrix is built by summing up the predictions of the 10 testing folds. During the fitting procedure, the class unbalancing is tackled by penalizing the misclassification of the minority class with a multiplicative factor inversely proportional to the class frequencies.

In order to evaluate the significance of the association between TLR7 mutations and COVID severity, the Fisher’s Exact Test was used.

For the quantitative PCR assay, the fold change in mRNA expression level per gene were compared between the individual patients and controls using an unpaired t test on the log-transformed fold changes. P values < .05 were considered statistically significant.

**In Vitro Peripheral Mononuclear Blood Cell Experiments**

PBMCs were isolated by Ficoll‐Hypaque density gradient centrifugation as previously described ^3^. 5×10^5^ PBMCs from COVID-19 patients and four unaffected male and female controls were stimulated for 4 hours with the TLR7 agonist imiquimod at 5 μg/mL or cell culture medium. Total RNA extraction was performed with RNAeasy Plus Mini kit and gDNA eliminator mini spin columns (Qiagen^®^, Hilden, Germany), following the manufacturer's instructions. First-strand cDNA was synthesized from total RNA using High Capacity cDNA Reverse Transcription Kit following the manufacturer's instructions (Thermo Fisher Scientific^®^, Waltham, Massachusetts, United States). The Advanced Universal SYBR Green Supermix (BioRad, Redmond, WA, United States) was used. All reactions were performed using the CFX96 Real-Time machine detection system (BioRad, Redmond, WA, United States) and each sample was amplified in duplicate. The following primers were used: *TLR7*, 5’-CATCAAGAGGCTGCAGATTAAA-3’, 5’-GAAAAGATGTTGTTGGCCTCA-3’; *IFNg*, 5’-TGACCAGAGCATCCAAAAGA-3’, 5’-CTCTTCGACCTCGAAACAGC-3’; *IRF7*, 5’-CCATCTTCGACTTCAGAGTCTTC-3’, 5’-TCTAGGTGCACTCGGCACAG-3’; *ISG15*, 5’-GACAAATGCGACGAACCTCT-3’, 5’-GAACAGGTCGTCCTGCACAC-3’; *HPRT1*, 5’-TGACACTGGCAAAACAATGCA-3’, 5’-GGTCCTTTTCACCAGCAAGCT-3’.
